# Enhancing dengue diagnosis and surveillance by integrating machine learning technologies with the NS1 rapid test kit

**DOI:** 10.64898/2026.05.05.26352445

**Authors:** Chun-Kai Hwang, Ying-Wen Chen, Yu-Tseng Wang, Tzong-Shiann Ho, Yen-Jen Oyang

## Abstract

**Background:** Dengue has been a major health threat globally in recent years. In particular, dengue incidences continue to increase annually and the epidemic area has expanded primarily due to global warming. Therefore, effective case detection and surveillance strategies are crucial to tackle this global health challenge. In clinical practice, the rapid test kit detecting dengue non-structural protein 1 antigen and commonly referred as NS1, is widely employed for early diagnosis. However, real-world studies revealed that the sensitivity of the NS1 test kit ranged from approximately 61% to 95%. Since early diagnosis is really critical for disease surveillance in the early stage of a dengue epidemic, scientists have been working hard to develop novel diagnosis methods that can provide higher sensitivity levels.

**Methodology/Principal Findings:** In response to this challenge, in this study, we have developed a novel diagnosis procedure that integrates machine learning technologies with the NS1 test kit. Our experimental results revealed that we would be able to raise the sensitivity of the dengue diagnosis procedure to higher than 99% by incorporating machine learning based prediction models to screen the suspected patients with a negative NS1 result. Furthermore, the relative risks between the suspected patients who were predicted to be positive and those who were predicted to be negative exceeded 4.8.

**Conclusions/Significance:** These results illustrate that the proposed approach provides an effective and efficient diagnosis procedure to address the global health challenge caused by spread of dengue.

**Author Summary:** This study has aimed to enhance surveillance of the dengue disease by integrating machine learning technologies with the rapid test kit commonly employed in early diagnosis. In clinical practice, the NS1 rapid test kit is widely employed for early diagnosis. However, real-world studies revealed that a certain percentage of the patients with a negative NS1 test result, ranging from 5% to 39%, were actually infected by dengue. Since early diagnosis is critical for disease control in the early stage of a dengue epidemic, scientists have been working hard to tackle this challenge. Based on this observation, this study was launched to investigate the effects of incorporating machine learning based prediction models to further screen those patients with a negative NS1 test result. The experimental results revealed that the proposed approach was able to identify over 99% of the patients who were infected by the dengue disease. Furthermore, the risk of the suspected patients who were predicted to be positive was 4.8 times higher than the risk of those who were predicted to be negative. The experimental results illustrate that the proposed approach provides an effective and efficient diagnosis procedure to enhance surveillance of the dengue disease.

## Introduction

Dengue is a mosquito-borne viral disease of major global public health importance, imposing a substantial and growing burden across tropical and subtropical regions worldwide. Recent estimates suggest that dengue causes between 100 and 400 million infections annually, with a marked increase in both incidence and geographic expansion over the past decade [1].

According to the World Health Organization and contemporary global modelling studies, dengue incidence has increased more than tenfold since the early 2000s, driven by climate change, rapid urbanization, population mobility, globalization, and persistent challenges in vector control [1]. In 2024 alone, over 14 million dengue cases and more than 12,000 dengue-related deaths were reported globally [1], underscoring the urgency of improving early case detection and surveillance strategies.

Clinically, dengue infection manifests across a wide spectrum, ranging from self-limited febrile illness to severe disease characterized by plasma leakage, hemorrhage, organ dysfunction, shock, and death [2]. Vulnerable populations—including older adults, young children, and patients with chronic comorbidities—are at particularly high risk of adverse outcomes [3]. The disease course typically progresses through febrile, critical, and recovery phases, with the critical phase often coinciding with defervescence and rapid clinical deterioration [4,5]. Importantly, early clinical features of dengue are nonspecific and frequently indistinguishable from other acute febrile illnesses, complicating timely diagnosis in emergency and outpatient settings [2,5].

Accurate and early laboratory diagnosis is therefore essential for appropriate clinical management, patient triage, and public health response. Among available diagnostic tools, detection of dengue non-structural protein 1 (NS1) antigen is widely used for early diagnosis, particularly within the first week of illness [6,7]. NS1 antigen tests offer operational advantages, including rapid turnaround and independence from host immune responses, and are thus routinely deployed in endemic regions and outbreak settings [8]. However, accumulating evidence demonstrates that NS1 test sensitivity is imperfect and context-dependent, influenced by factors such as viral serotype, timing of specimen collection, host immune status, and secondary infection [9–12]. Meta-analyses and real-world studies report NS1 sensitivity ranging from approximately 61% to 95% and a specificity above 90% [7,12,13], implying that a clinically meaningful proportion of true dengue cases remain undetected when NS1 is used as a standalone diagnostic tool.

From a public-health perspective, these NS1-negative dengue cases represent a critical blind spot. Missed diagnoses not only delay appropriate clinical care but also undermine outbreak detection, epidemiological surveillance, and timely vector-control interventions. Consequently, there is increasing interest in complementary diagnostic strategies that leverage routinely available clinical data to enhance early dengue detection, particularly among NS1-negative patients.

Machine learning (ML) approaches have emerged as promising tools for addressing complex diagnostic challenges in infectious diseases. By integrating multidimensional clinical, demographic, and laboratory data, ML models can capture nonlinear relationships and interaction effects that may not be apparent using conventional statistical methods. Prior studies have demonstrated the potential of ML-based models to support dengue diagnosis, severity prediction, and triage decision-making [14–33]. However, most existing work has focused on general dengue prediction rather than the specific and clinically important subgroup of NS1-negative cases.

In this study, we have aimed to address this unmet need by developing and validating machine learning-based prediction models specifically designed to identify dengue infection among patients with a negative NS1 antigen test result. Fig 1 illustrates the approach proposed in this study. The machine learning-based prediction model is incorporated to screen patients with a negative NS1 antigen test result. Based on a large emergency-department cohort from a major tertiary medical center in Taiwan, we investigated the effects of incorporating four alternative ML algorithms—logistic regression (LR), classification and regression tree (CART), artificial neural networks (ANN), and support vector machines (SVM)—with rigorous feature selection and class-imbalance handling. Our overarching objective has been to illustrate how ML-assisted screening can meaningfully enhance diagnostic sensitivity when integrated with routine NS1 testing, thereby strengthening early clinical recognition and outbreak surveillance.

**Fig 1.**
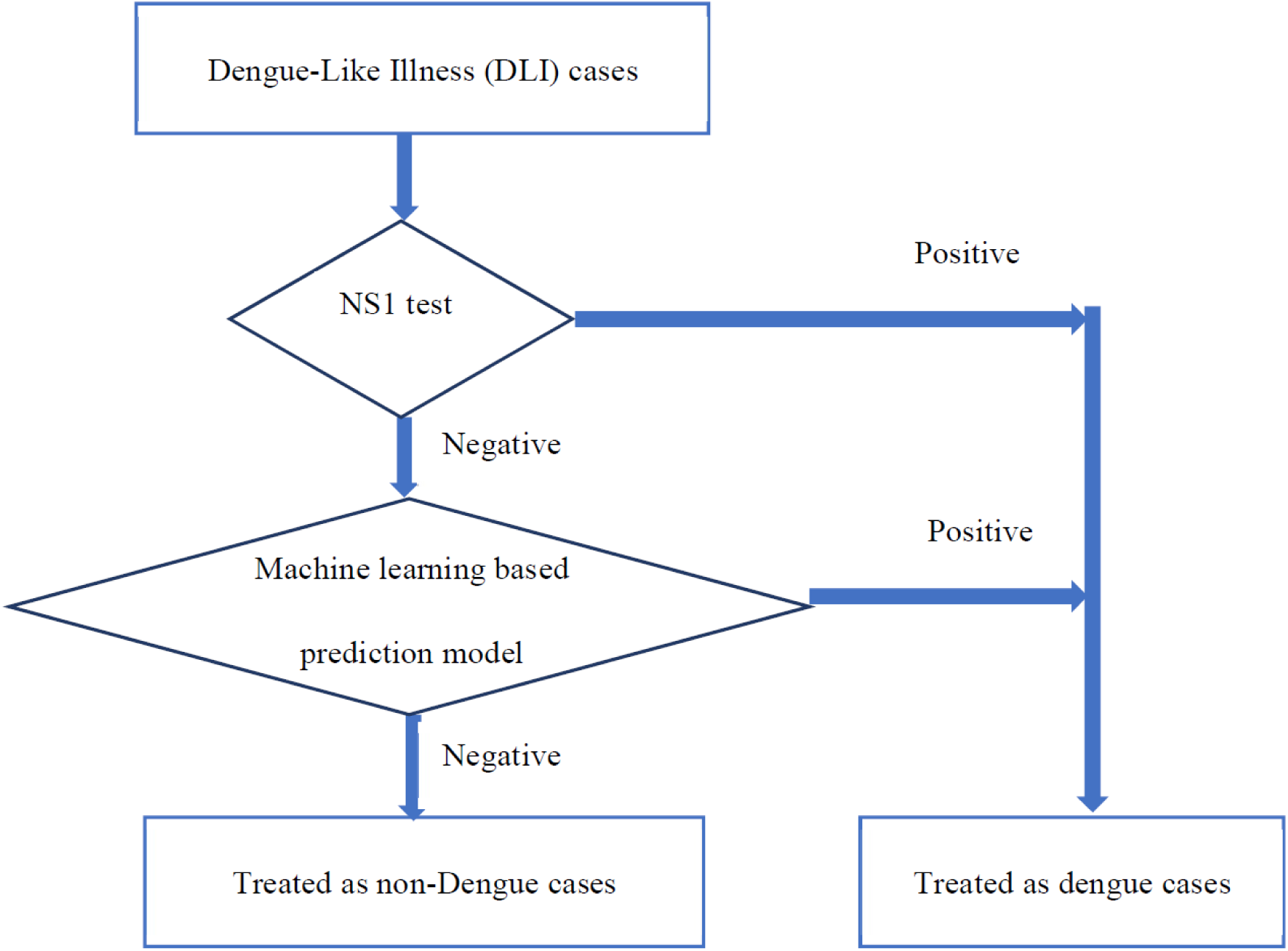
The proposed procedure that integrates the NS1 test and the machine learning-based prediction model for dengue diagnosis.

## Methods

### Data collection and outcome measurement

Fig 2 depicts the data collection and processing workflow for this study. In 2015, a total of 100,491 patients were admitted to the emergency department (ED) at National Cheng Kung University Hospital (NCKUH). Among these patients, 3,698 patients who canceled their ED consultation and 6,611 patients who were readmitted within 36 hours were excluded from the analysis. In the remaining cohort, 6,368 patients met our definition of a dengue-like illness (DLI) case, given that (1) the patient was coded with ICD-9 061 (dengue), 0654 (mosquito-borne hemorrhagic fever), 0663 (other mosquito-borne fever), or v735 (screening examination for other arthropod-borne viral diseases) for dengue fever; or (2) the patient received one or more dengue serological and/or virological tests, including dengue NS1, dengue-IgM, viral load of DENV, or dengue serotyping using polymerase chain reaction (PCR) to detect DENV-1 and DENV-2. We then excluded 1,474 DLI cases due to missing laboratory data or incomplete information in key variables. In the end, the DLI dataset employed in this study comprised 4,894 cases, among which 2,942 were confirmed dengue cases and 1,952 were Non-Dengue cases. The demographic analysis of these 4,894 cases is presented in Table 1.

**Fig 2.**
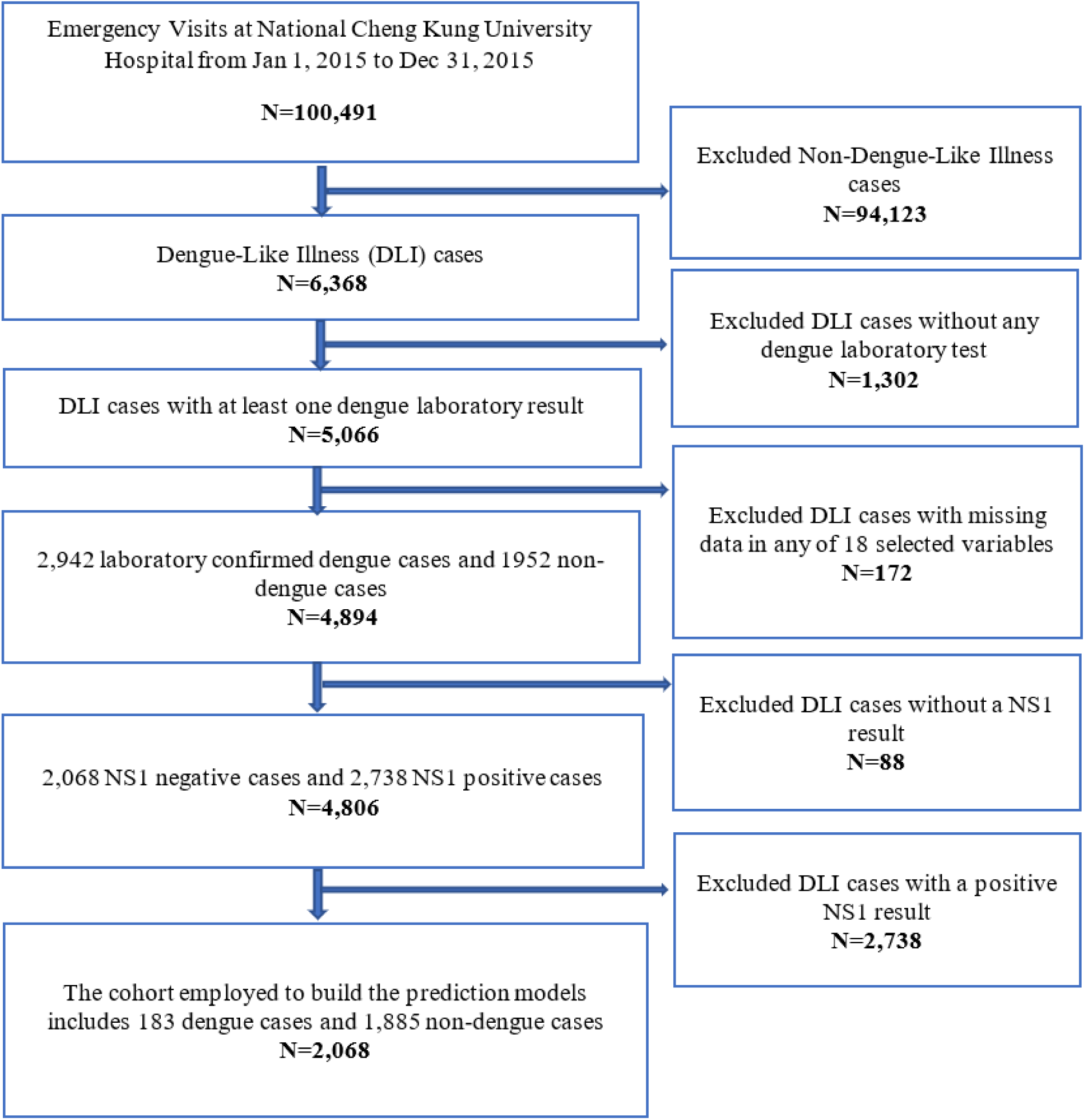
Flow diagram for extracting 2,068 NS1-negative cases with 183 laboratory-confirmed dengue (case group) and 1,885 Non-Dengue cases (control group).

**Table 1.**
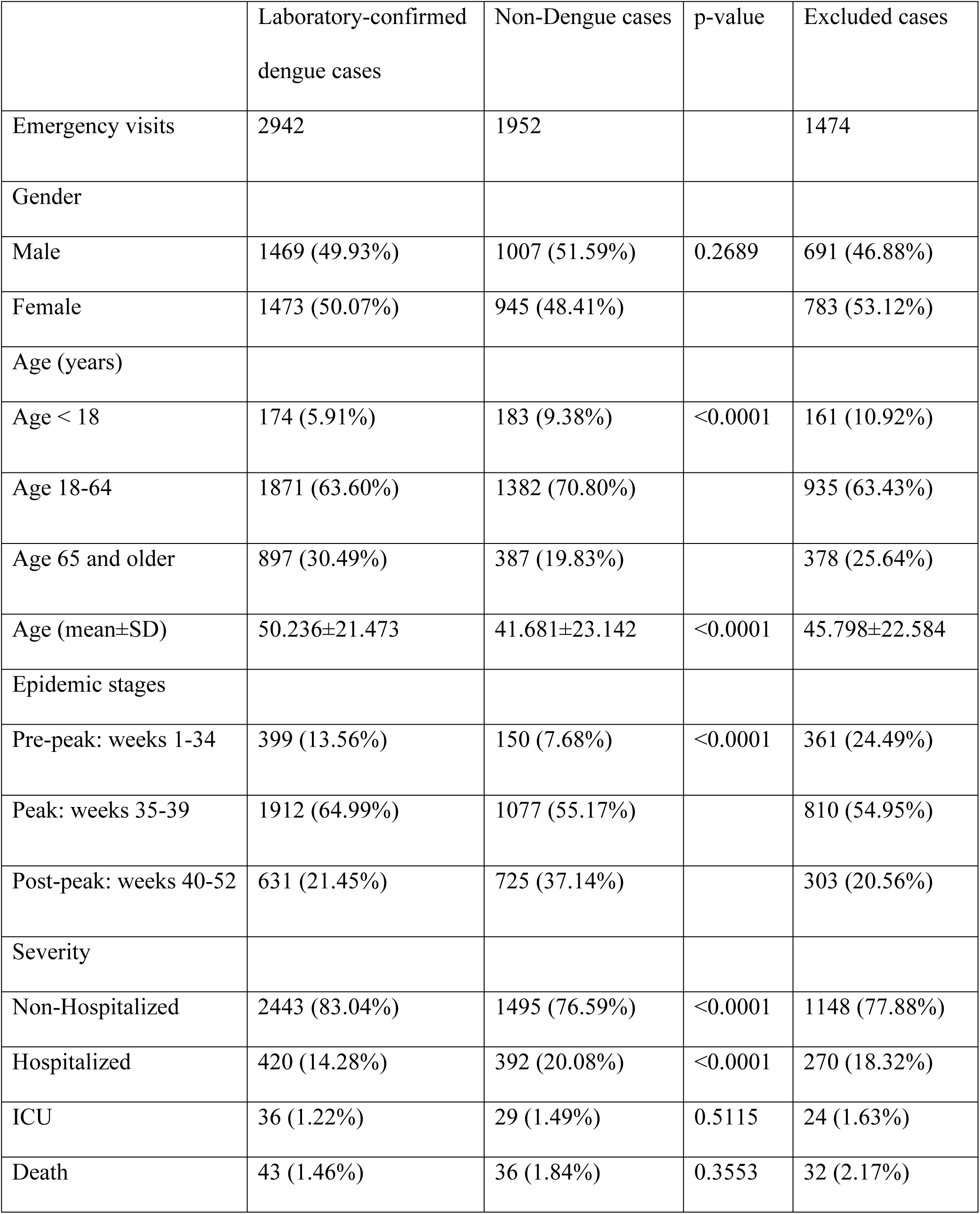
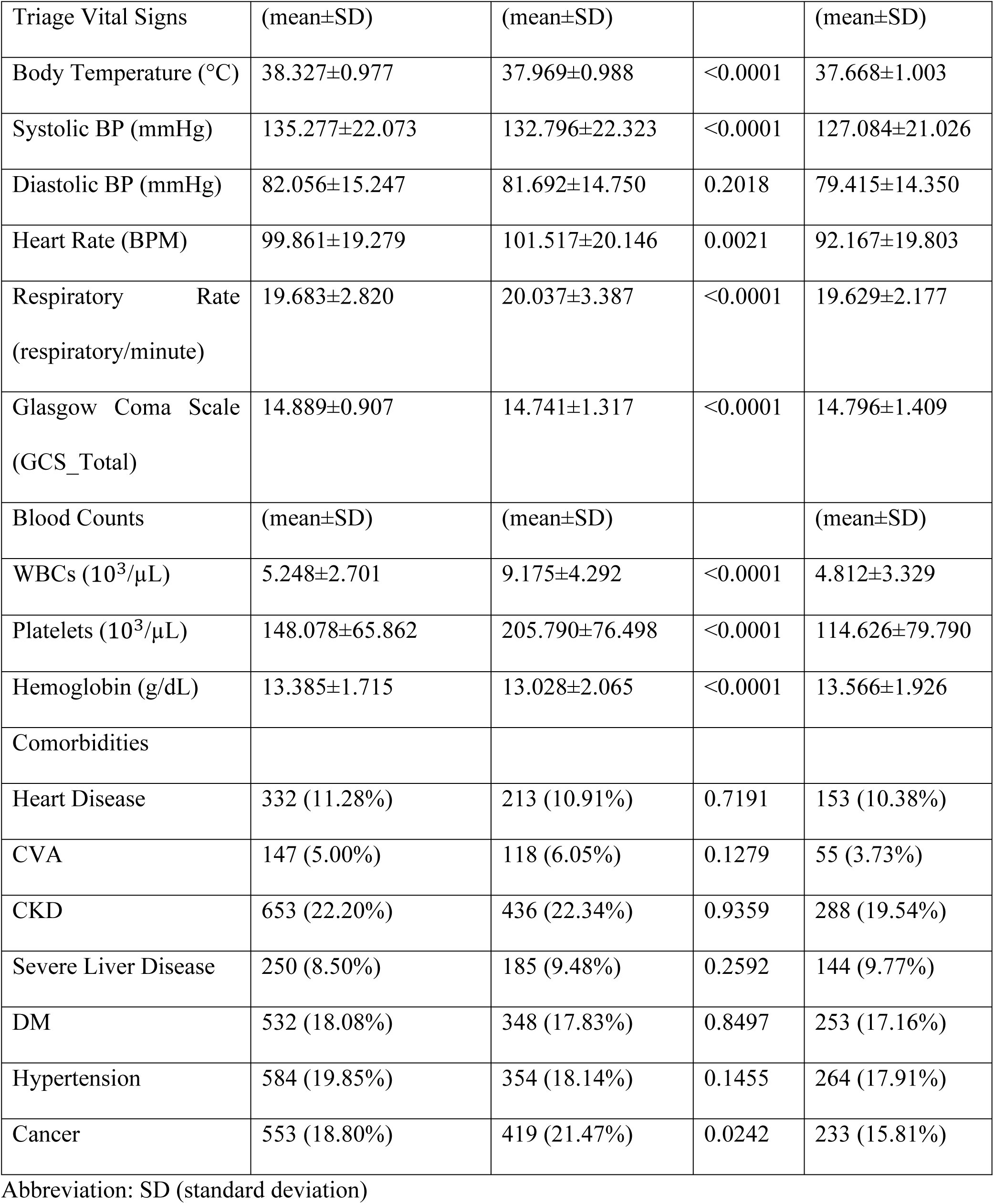
Demographic analysis of the 4,894 DLI cases.

|  | Laboratory-confirmed<br>dengue cases | Non-Dengue cases | p-value | Excluded cases |
| --- | --- | --- | --- | --- |
| Emergency visits | 2942 | 1952 |  | 1474 |
| Gender |  |  |  |  |
| Male | 1469 (49.93%) | 1007 (51.59%) | 0.2689 | 691 (46.88%) |
| Female | 1473 (50.07%) | 945 (48.41%) |  | 783 (53.12%) |
| Age (years) |  |  |  |  |
| Age < 18 | 174 (5.91%) | 183 (9.38%) | <0.0001 | 161 (10.92%) |
| Age 18-64 | 1871 (63.60%) | 1382 (70.80%) |  | 935 (63.43%) |
| Age 65 and older | 897 (30.49%) | 387 (19.83%) |  | 378 (25.64%) |
| Age (mean±SD) | 50.236±21.473 | 41.681±23.142 | <0.0001 | 45.798±22.584 |
| Epidemic stages |  |  |  |  |
| Pre-peak: weeks 1-34 | 399 (13.56%) | 150 (7.68%) | <0.0001 | 361 (24.49%) |
| Peak: weeks 35-39 | 1912 (64.99%) | 1077 (55.17%) |  | 810 (54.95%) |
| Post-peak: weeks 40-52 | 631 (21.45%) | 725 (37.14%) |  | 303 (20.56%) |
| Severity |  |  |  |  |
| Non-Hospitalized | 2443 (83.04%) | 1495 (76.59%) | <0.0001 | 1148 (77.88%) |
| Hospitalized | 420 (14.28%) | 392 (20.08%) | <0.0001 | 270 (18.32%) |
| ICU | 36 (1.22%) | 29 (1.49%) | 0.5115 | 24 (1.63%) |
| Death | 43 (1.46%) | 36 (1.84%) | 0.3553 | 32 (2.17%) |

| Triage Vital Signs | (mean±SD) | (mean±SD) |  | (mean±SD) |
| --- | --- | --- | --- | --- |
| Body Temperature (°C) | 38.327±0.977 | 37.969±0.988 | <0.0001 | 37.668±1.003 |
| Systolic BP (mmHg) | 135.277±22.073 | 132.796±22.323 | <0.0001 | 127.084±21.026 |
| Diastolic BP (mmHg) | 82.056±15.247 | 81.692±14.750 | 0.2018 | 79.415±14.350 |
| Heart Rate (BPM) | 99.861±19.279 | 101.517±20.146 | 0.0021 | 92.167±19.803 |
| Respiratory Rate<br>(respiratory/minute) | 19.683±2.820 | 20.037±3.387 | <0.0001 | 19.629±2.177 |
| Glasgow Coma Scale<br>(GCS_Total) | 14.889±0.907 | 14.741±1.317 | <0.0001 | 14.796±1.409 |
| Blood Counts | (mean±SD) | (mean±SD) |  | (mean±SD) |
| WBCs (10<sup>3</sup>/μL) | 5.248±2.701 | 9.175±4.292 | <0.0001 | 4.812±3.329 |
| Platelets (10<sup>3</sup>/μL) | 148.078±65.862 | 205.790±76.498 | <0.0001 | 114.626±79.790 |
| Hemoglobin (g/dL) | 13.385±1.715 | 13.028±2.065 | <0.0001 | 13.566±1.926 |
| Comorbidities |  |  |  |  |
| Heart Disease | 332 (11.28%) | 213 (10.91%) | 0.7191 | 153 (10.38%) |
| CVA | 147 (5.00%) | 118 (6.05%) | 0.1279 | 55 (3.73%) |
| CKD | 653 (22.20%) | 436 (22.34%) | 0.9359 | 288 (19.54%) |
| Severe Liver Disease | 250 (8.50%) | 185 (9.48%) | 0.2592 | 144 (9.77%) |
| DM | 532 (18.08%) | 348 (17.83%) | 0.8497 | 253 (17.16%) |
| Hypertension | 584 (19.85%) | 354 (18.14%) | 0.1455 | 264 (17.91%) |
| Cancer | 553 (18.80%) | 419 (21.47%) | 0.0242 | 233 (15.81%) |

Abbreviation: SD (standard deviation)

As mentioned earlier, the primary objective of this study has been to develop machine learning-based prediction models that can identify dengue cases who had a negative NS1 result. Therefore, we extracted the 2,068 DLI cases with a negative NS1 result, i.e., excluding the 88 DLI cases without an NS1 result and the 2,738 DLI cases with a positive NS1 result, to build our prediction models. Among the 2,068 cases, 1,885 were Non-Dengue cases and 183 were dengue cases, which were confirmed by other laboratory results. Table 2 shows the demographic analysis of these 2,068 cases.

**Table 2.** Demographic analysis of the cohort employed to build prediction models in this study.

|  | Dengue cases with a negative NS1 result | Non-Dengue cases with a negative NS1 result | p-value | Excluded cases with a positive NS1 result |
| --- | --- | --- | --- | --- |
| Emergency visits | 183 | 1885 |  | 2738 |
| Gender |  |  |  |  |
| Male | 107 (58.47%) | 971 (51.51%) | 0.0852 | 1350 (49.31%) |
| Female | 76 (41.53%) | 914 (48.49%) |  | 1388 (50.69%) |
| Age (years) |  |  |  |  |
| Age < 18 | 4 (2.19%) | 159 (8.44%) | <0.0001 | 154 (5.62%) |
| Age 18-64 | 110 (60.11%) | 1355 (71.88%) |  | 1758 (64.21%) |
| Age 65 and older | 69 (37.70%) | 371 (19.68%) |  | 826 (30.17%) |
| Age (mean±SD) | 55.607±21.144 | 41.798±22.968 | <0.0001 | 50.093±21.333 |
| Epidemic stages |  |  |  |  |
| Pre-peak: weeks 1-34 | 26 (14.21%) | 148 (7.85%) | 0.0122 | 371 (13.55%) |
| Peak: weeks 35-39 | 94 (51.37%) | 1020 (54.11%) |  | 1809 (66.07%) |
| Post-peak: weeks 40-52 | 63 (34.43%) | 717 (38.04%) |  | 558 (20.38%) |
| Severity |  |  |  |  |
| Non-Hospitalized | 126 (68.85%) | 1450 (76.92%) | 0.0184 | 2304 (84.15%) |
| Hospitalized | 43 (23.50%) | 374 (19.84%) | 0.2799 | 371 (13.55%) |
| ICU | 11 (6.01%) | 27 (1.43%) | <0.0001 | 23 (0.84%) |
| Death | 3 (1.64%) | 34 (1.80%) | 1 | 40 (1.46%) |
| Triage Vital Signs | (mean±SD) | (mean±SD) |  | (mean±SD) |
| Body Temperature (°C) | 37.685±1.031 | 37.958±0.988 | 0.0004 | 38.376±0.955 |
| Systolic BP (mmHg) | 140.104±23.451 | 132.889±22.251 | <0.0001 | 135.118±21.883 |
| Diastolic BP (mmHg) | 84.011±15.135 | 81.822±14.635 | 0.0312 | 81.977±15.254 |
| Heart Rate (BPM) | 96.809±18.998 | 101.428±20.133 | 0.001 | 100.103±19.289 |
| Respiratory Rate<br>(respiratory/minute) | 20.077±5.224 | 20.028±3.417 | 0.4507 | 19.650±2.581 |
| Glasgow Coma Scale<br>(GCS_Total) | 14.710±1.526 | 14.737±1.334 | 0.4087 | 14.903±0.841 |
| Blood Counts | (mean±SD) | (mean±SD) |  | (mean±SD) |
| WBCs (10<sup>3</sup>/μL) | 7.599±4.696 | 9.268±4.277 | <0.0001 | 5.100±2.431 |
| Platelets (10<sup>3</sup>/μL) | 161.131±89.688 | 206.871±76.326 | <0.0001 | 147.497±63.897 |
| Hemoglobin (g/dL) | 13.069±2.413 | 13.027±2.081 | 0.409 | 13.396±1.650 |
| Comorbidities |  |  |  |  |
| Heart Disease | 20 (10.93%) | 211 (11.19%) | 1 | 312 (11.40%) |
| CVA | 9 (4.92%) | 106 (5.62%) | 0.8192 | 136 (4.97%) |
| CKD | 51 (27.87%) | 427 (22.65%) | 0.132 | 600 (21.91%) |
| Severe Liver Disease | 12 (6.56%) | 183 (9.71%) | 0.2077 | 236 (8.62%) |
| DM | 36 (19.67%) | 334 (17.72%) | 0.5774 | 494 (18.04%) |
| Hypertension | 28 (15.30%) | 340 (18.04%) | 0.4106 | 552 (20.16%) |
| Cancer | 43 (23.50%) | 406 (21.54%) | 0.6033 | 506 (18.48%) |

Abbreviation: SD (standard deviation)

### Feature selection

It is well known that inclusion of irrelevant variables in machine learning models can introduce noise, diminish predictive performance, and reduce the interpretability of model outputs [34]. Furthermore, unnecessary variables increase computational complexity, resulting in higher processing times and memory usage during model training. Therefore, a common practice before the machine learning models are built is to conduct feature selection.

#### Feature selection with blood examination data

For building the prediction models that operated with the blood examination data available, the following 18 variables were initially included: age, gender, systolic blood pressure (SBP), diastolic blood pressure (DBP), body temperature, heart rate (beats per minute, BPM), respiratory rate, Glasgow Coma Scale total score (GCS_Total), heart disease, hypertension, diabetes mellitus (DM), cancer, chronic kidney disease (CKD), severe liver disease, cerebrovascular accident (CVA), platelet count (exam_Plt), hemoglobin concentration (exam_Hb), and white blood cell count (exam_WBC). Then, three multivariate analysis methods, namely the logistic regression [35], the least absolute shrinkage and selection operator (LASSO) [36] and the ensemble variant of the minimum redundancy maximum relevance (mRMR) [37], were employed to identify 6 important variables. Table 3 shows the 6 important variables identified by alternative methods and the union of the identified variables. The union, which included 9 variables, and the set of initial 18 variables were then employed to build the prediction models.

**Table 3.** The feature sets identified by mRMR, LR, LASSO with the blood examination data incorporated.

| Feature selection | 6 variables by mRMR | 6 variables by LASSO | 6 variables by LR | Union of 9 variables |
| --- | --- | --- | --- | --- |
| Feature set | Age<br>exam_WBC<br>DBP<br>SevereLiverDisease<br>Temperature<br>CVA | Temperature<br>exam_WBC<br>Age<br>exam_Plt<br>SBP<br>exam_Hb | Age<br>exam_Plt<br>exam_WBC<br>Temperature<br>SevereLiverDisease<br>exam_Hb | Age<br>exam_WBC<br>DBP<br>SevereLiverDisease<br>Temperature<br>CVA<br>exam_Plt<br>SBP<br>exam_Hb |

#### Feature selection without blood examination data

For building the prediction models that operated without the blood examination data available, the following 15 clinical and demographic variables were initially included: age, gender, SBP, DBP, body temperature, BPM, respiratory rate, GCS_Total, heart disease, hypertension, DM, cancer, CKD, severe liver disease, and CVA. We then followed the same procedure as described above to generate the feature sets shown in Table 4. The union, which included 9 variables, and the set of initial 15 variables were then employed to build the prediction models.

**Table 4.** The feature sets identified by mRMR, LR, LASSO without blood examination data incorporated.

| Feature selection | 6 variables by mRMR | 6 variables by LASSO | 6 variables by LR | Union of 9 variables |
| --- | --- | --- | --- | --- |
| Feature set | Age<br>Temperature<br>Gender<br>SevereLiverDisease<br>DBP<br>CVA | Temperature<br>Gender<br>Hypertension<br>SevereLiverDisease<br>Age<br>SBP | Age<br>Temperature<br>Gender<br>Hypertension<br>SevereLiverDisease<br>GCS_Total | Age<br>Temperature<br>Gender<br>SevereLiverD<br>isease<br>DBP<br>CVA<br>Hypertension<br>SBP<br>GCS_Total |

### Experimental procedure to build and evaluate the prediction models

Fig 3 illustrates the experimental procedure to build and evaluate the prediction models. In this study, four widely utilized machine learning algorithms—the logistic regression (LR), classification and regression tree (CART) [38], artificial neural networks (ANN) [21], and support vector machines (SVM) [39]—were employed to develop prediction models. Since the dataset exhibited significant class imbalance, consisting of 183 dengue cases and 1,885 Non-Dengue cases, the Random Over-Sampling Examples (ROSE) method [40,41] was employed to generate a balanced training dataset.

**Fig 3.**
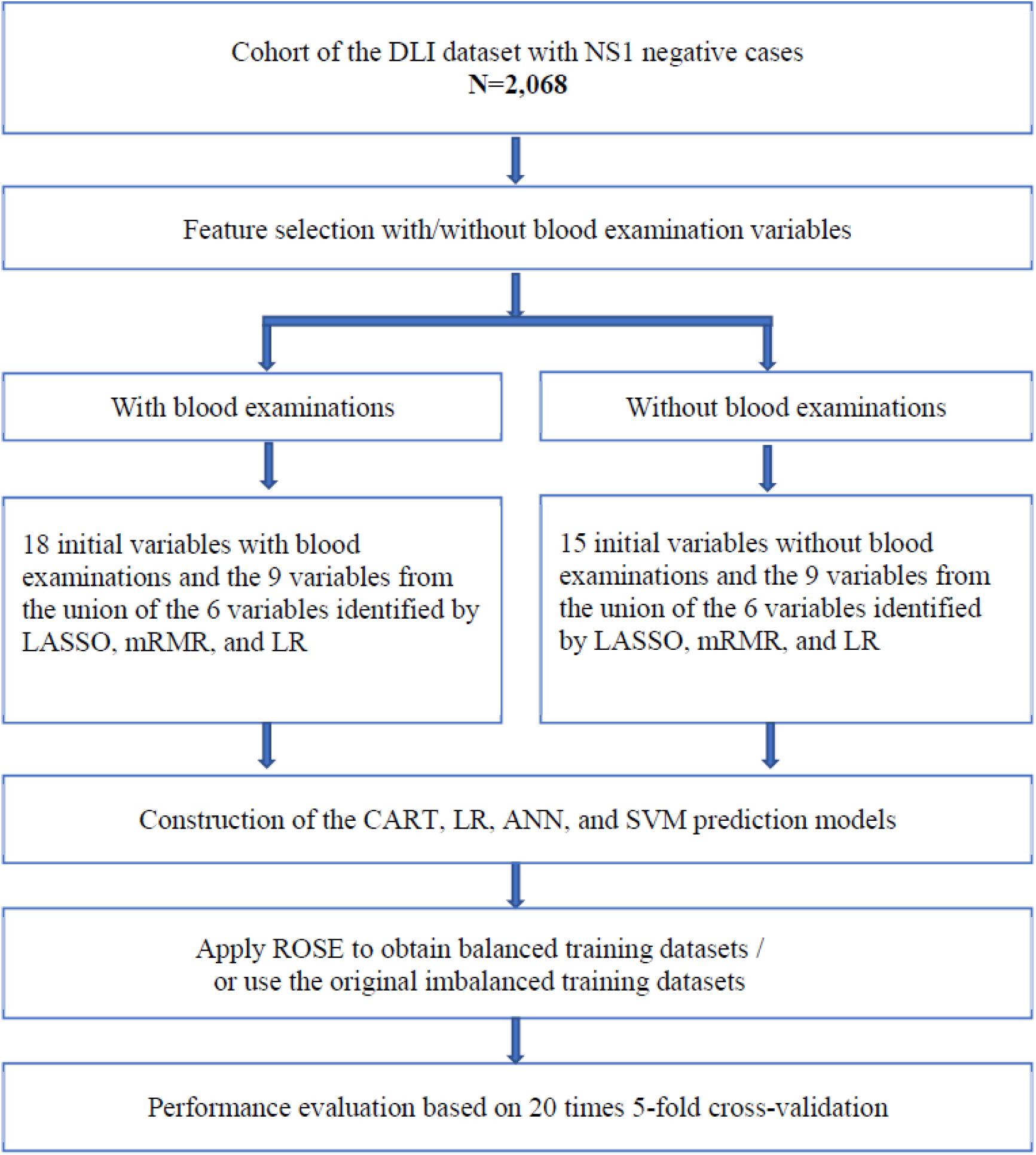
Experimental procedure to build and evaluate the prediction models.

Model performance was evaluated through 20 iterations of 5-fold cross-validation to ensure statistical robustness and generalizability of the findings. Comprehensive analyses were then conducted by calibrating the models to achieve the following sensitivity levels: 70% and 85%.

The parameter settings for each machine learning algorithm, along with details of the computational environment and software libraries, are summarized in Table 5. Optimal hyperparameter settings for the SVM and multilayer ANN models were determined based on grid search with 5-fold cross-validation. This rigorous approach ensured the development of highly tuned and robust prediction models.

**Table 5.** The packages and parameter combinations evaluated for building LR, CART, ANN, and SVM models.

| Model | Programming language | Package | Parameter settings |
| --- | --- | --- | --- |
| LR | Python version 3.7.3 | sklearn.linear_model<br>LogisticRegression | solver='lbfgs',<br>penalty='l2',<br>max_iter=100 |
| CART | R version 4.3.1 | rpart | split = "information",<br>prior=0.1~0.9 with 0.008 step size,<br>cp=[0.008, 0.007, 0.006, 0.005, 0.004,<br>0.003, 0.002, 0.001, 0.0009, 0.0008,<br>0.0007, 0.0006] |
| ANN | Python version 3.7.3 | TensorFlow<br>Keras | Input neurons = [4, 6, 9] for 4-, 6-, or<br>9-feature sets, |
|  |  |  | Hidden layers = [3, 4, 5]<br>Hidden neurons = [12, 16, 20] |
| SVM | Python version 3.7.3 | sklearn<br>svm | kernel=['linear','rbf','poly'],<br>C=[0.01, 0.1, 1,3, 10, 15,<br>20, 25, 40, 50, 75, 100],<br>gamma='auto',<br>probability=True |

### Ethics statement

The study was conducted in accordance with the principles of the Declaration of Helsinki. The study protocols were approved by the Institutional Review Boards of National Cheng Kung University Hospital (NCKUH) (reference number A-ER-114-331), which is organized and operated according to the laws and regulations of Good Clinical Practice (ICH-GCP). All clinical data were fully de-identified and anonymized to protect individual privacy, and only aggregated data were used for further analyses and statistical tests. Therefore, individual consent was waived in the current study.

## Results

### Performance of the prediction models

Table 6 presents how the alternative prediction models built with blood examination variables incorporated performed in terms of the area under the receiver operating characteristic curve, which is commonly referred to as the AUC. Meanwhile, Table 7 presents how the alternative prediction models built without blood examination variables incorporated performed. As mentioned earlier, in this study, performance was evaluated through 20 iterations of 5-fold cross-validation. A general observation is that the prediction models built with the feature sets identified by the feature selection processes marginally outperformed the prediction models built with the initial feature sets in terms of AUCs. Furthermore, it is observed that the LR models marginally outperformed alternative prediction models and the LR models built with blood examination variables incorporated marginally outperformed the LR models built without blood examination variables incorporated. Accordingly, in the following discussion, we will focus on the effects of incorporating the LR models to enhance disease diagnosis as shown in Fig 1.

**Table 6.** AUCs, in terms of mean (95% confidence interval), of the alternative prediction models built with blood examination variables incorporated.

| ML model | Feature set | Model with sensitivity level 70% | Model with sensitivity level 85% |
| --- | --- | --- | --- |
| CART | 18 features | 0.682 (0.675 - 0.688) | 0.682 (0.674 - 0.691) |
|  | 9 features | 0.689 (0.681 - 0.697) | 0.685 (0.677 - 0.693) |
| LR | 18 features | 0.708 (0.705 - 0.712) | 0.708 (0.705 - 0.712) |
|  | 9 features | 0.721 (0.718 - 0.723) | 0.721 (0.718 - 0.723) |
| ANN | 18 features | 0.634 (0.617 - 0.650) | 0.634 (0.617 - 0.650) |
|  | 9 features union | 0.672 (0.658 - 0.686) | 0.672 (0.658 - 0.686) |
| SVM | 18 features | 0.697 (0.692 - 0.702) | 0.697 (0.692 - 0.702) |
|  | 9 features union | 0.717 (0.715 - 0.719) | 0.717 (0.715 - 0.719) |

**Table 7.** AUCs, in terms of mean (95% confidence interval), of the alternative prediction models built without blood examination variables incorporated.

| ML model | Feature set | Model with sensitivity level 70% | Model with sensitivity level 85% |
| --- | --- | --- | --- |
| CART | 15 features | 0.638 (0.632 - 0.643) | 0.641 (0.634 - 0.648) |
|  | 9 features | 0.646 (0.640 - 0.651) | 0.648 (0.642 - 0.654) |
| LR | 15 features | 0.662 (0.658 - 0.667) | 0.662 (0.658 - 0.667) |
|  | 9 features | 0.678 (0.676 - 0.681) | 0.678 (0.676 - 0.681) |
| ANN | 15 features | 0.589 (0.572 - 0.606) | 0.589 (0.572 - 0.606) |
|  | 9 features | 0.569 (0.543 - 0.596) | 0.569 (0.543 - 0.596) |
| SVM | 15 features | 0.661 (0.657 - 0.666) | 0.661 (0.657 - 0.666) |
|  | 9 features | 0.678 (0.674 - 0.682) | 0.678 (0.674 - 0.682) |

Table 8 shows the overall sensitivities of the diagnosis procedure exhibited in Fig 1 when alternative LR models built with blood examination variables were incorporated. In this respect, the detailed performance data of the LR models, which include the positive predictive value (PPV), sensitivity, specificity, negative predictive value (NPV), correspond to what was observed in the experimental procedure exhibited in Fig 3. Furthermore, it must be noted that the sensitivity level of the NS1 test observed with our dataset, which was 93.7%, is in conformity with the sensitivity levels reported in other studies [7,12,13]. The performance data in Table 8 reveal that the overall sensitivity of the diagnosis procedure shown in Fig 1 reached 98% when the LR model with a sensitivity level at 70% was incorporated. Furthermore, the overall sensitivity reached 99% when the LR model with a sensitivity level at 85% was incorporated.

**Table 8.**
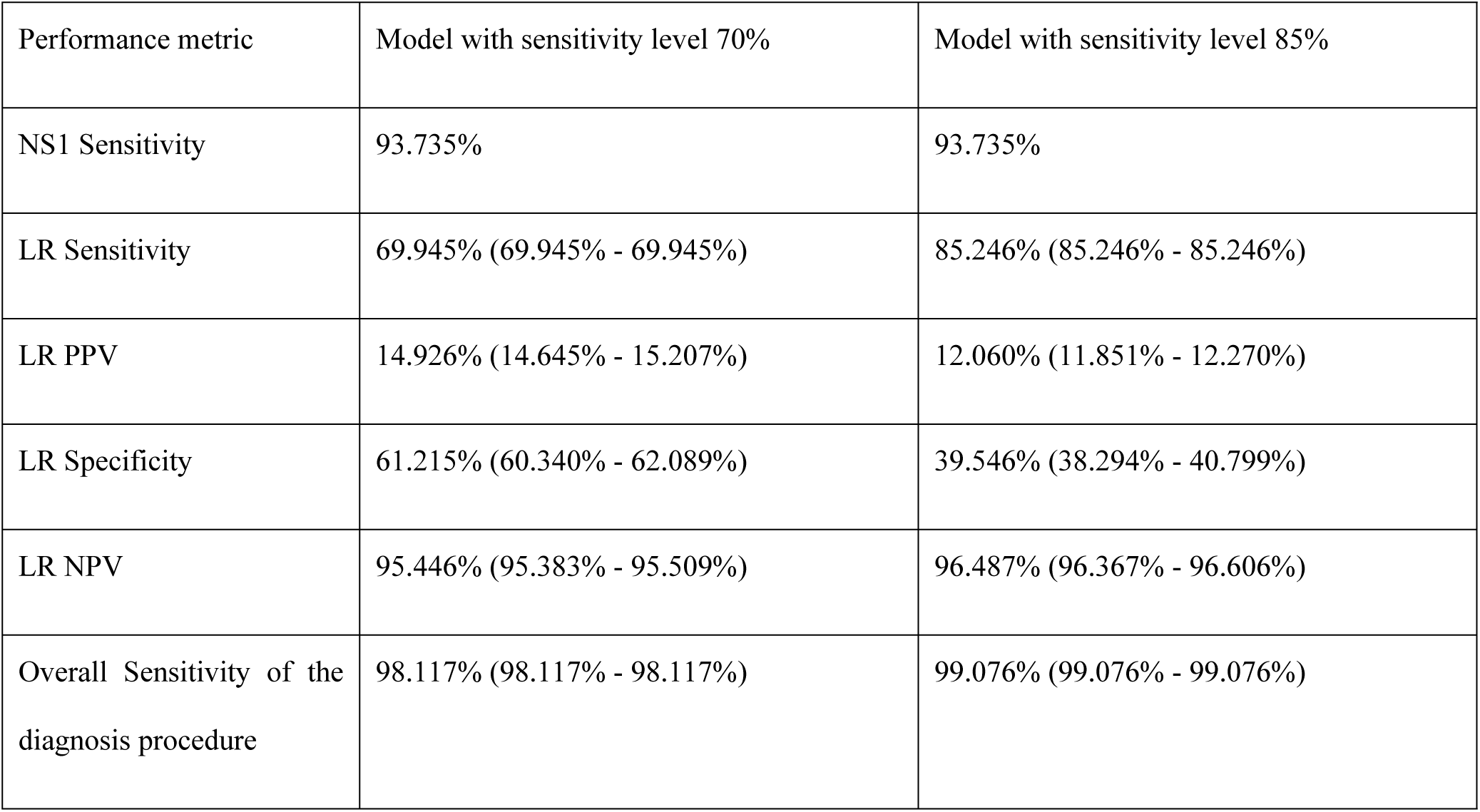
Overall effects of the diagnosis procedure shown in Fig 1 when alternative LR models built with blood examination variables were incorporated. The performance data are reported in terms of mean (95% confidence interval).

Table 9 shows how the LR models built with blood examination variables performed in different stages of the epidemic. The performance data reported include the relative risk (RR) between those patients that were predicted to be positive and those predicted to be negative and the F1 score, which is the harmonic mean of the PPV and the sensitivity and is widely employed in the machine learning research community to evaluate the overall performance of a prediction model, in addition to the PPV, sensitivity, specificity, and NPV. Since the primary objective of this study has been to develop a novel approach to enhance disease diagnosis in the early stage of a dengue epidemic, we will focus on the performance data in the pre-peak stage. The pre-peak, peak, and post-peak stages include weeks 1-34, weeks 35-39, and weeks 40-52 of 2015. The most important observation is that the LR model with a sensitivity level at 85% delivered significantly higher RR than the LR model with a sensitivity level at 70% in the pre-peak stage. This significant discrepancy was due to the substantial difference between their sensitivity levels, 89.423% vs. 62.500% in the pre-peak stage. Another important observation is that both of the two LR models built with blood examination variables delivered significantly higher PPV levels in the pre-peak stage than in the later stages of the epidemic.

**Table 9.**
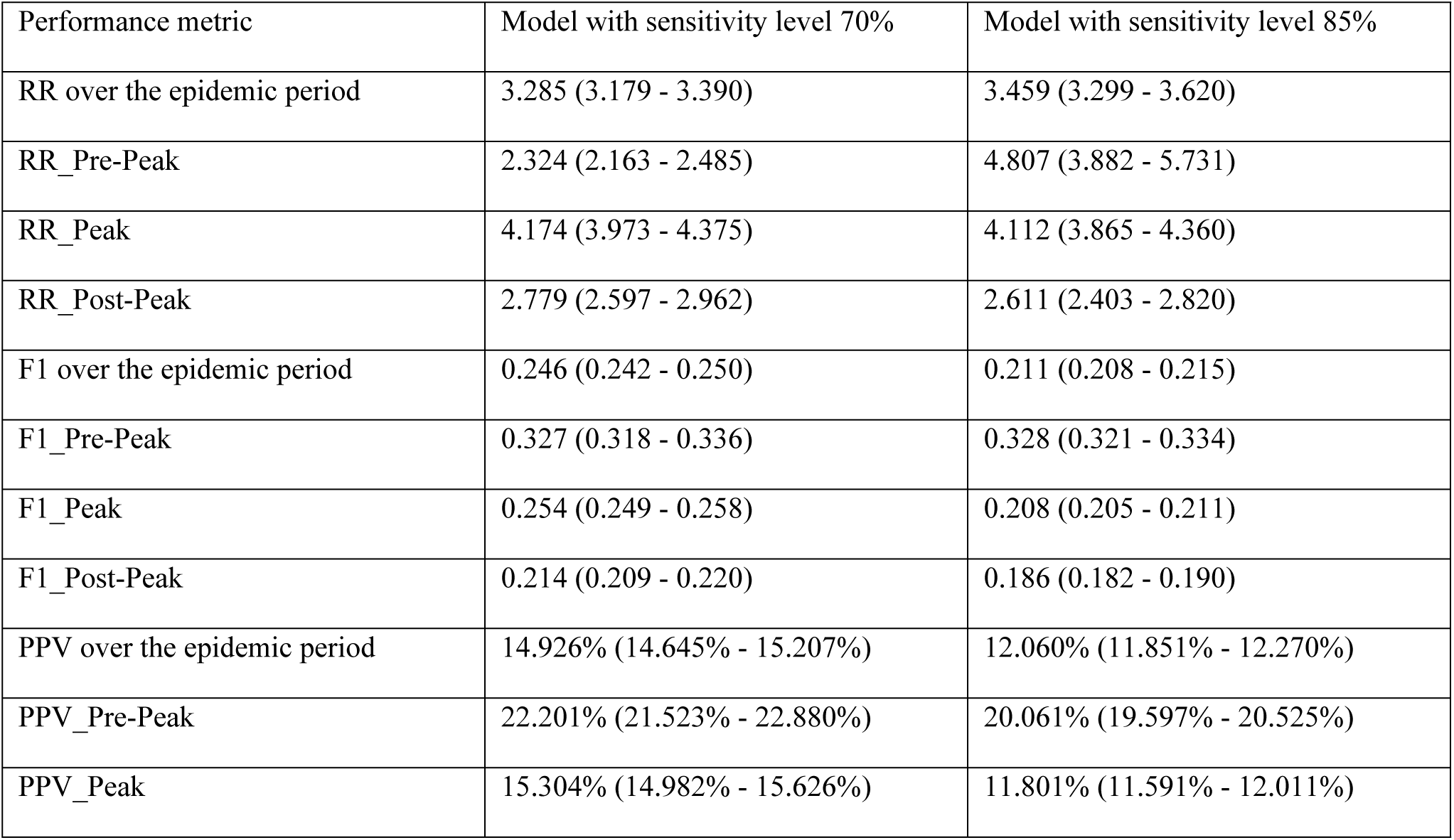
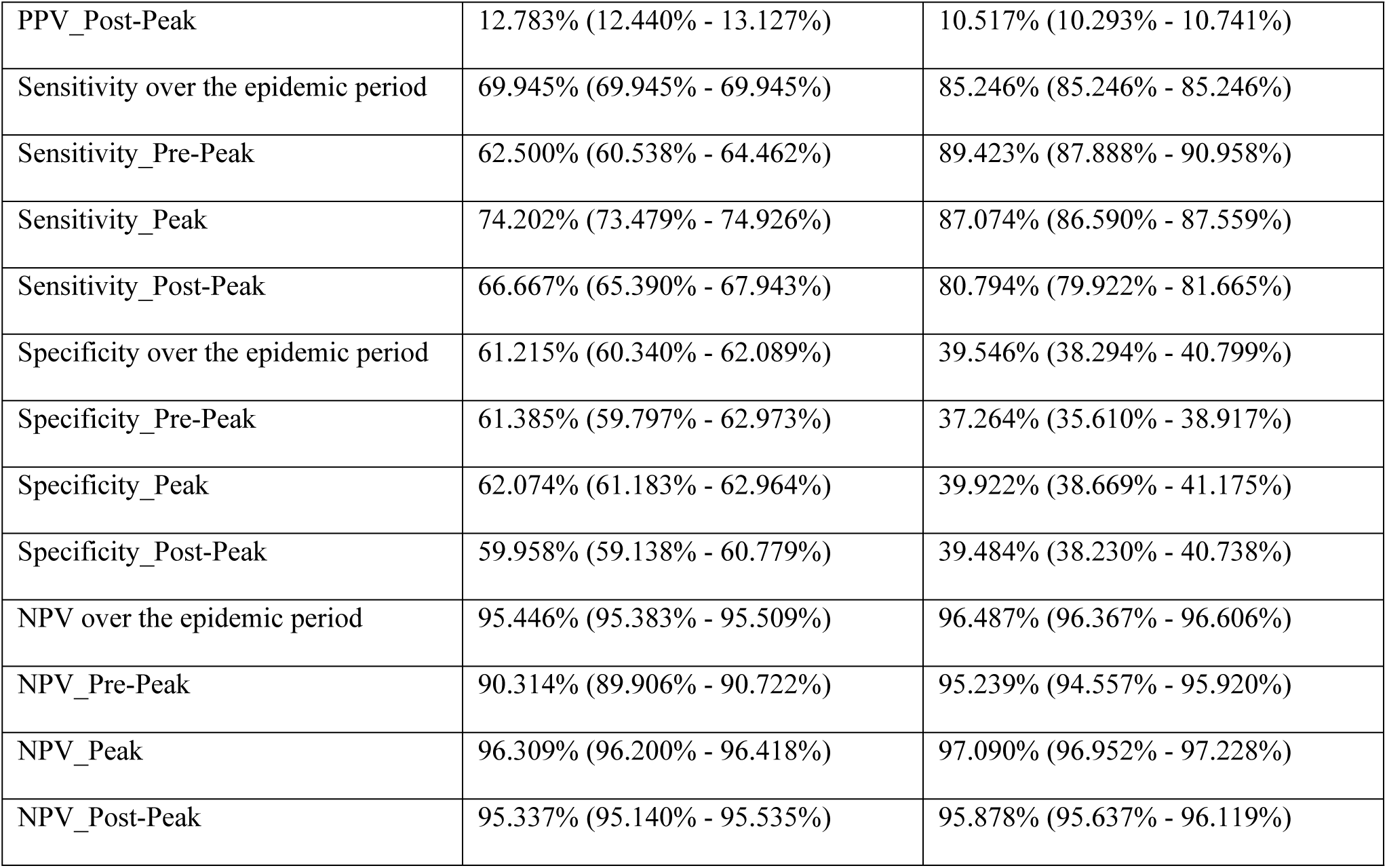
How the LR models built with blood examination variables performed in different stages of the epidemic. The performance data are reported in terms of mean (95% confidence interval).

Tables 10 and 11 show the performance data observed when the LR models built without blood examination variables were incorporated. As shown in Table 10, the overall sensitivities of the diagnosis procedure exhibited in Fig 1 reached 98% and 99% when the LR model with a sensitivity level at 70% and the LR model with a sensitivity level at 85% were incorporated, respectively. If we compare the detailed performance data in Tables 8 and 10, we can observe that the LR models built with blood examination variables marginally outperformed the LR models built without blood examination variables with respect to PPV and specificity. Furthermore, the LR models built with blood examination variables incorporated delivered significantly higher sensitivity levels than the LR models built without blood examination variables incorporated in the pre-peak stage. With respect to the PPV levels, the LR models built with blood examination variables incorporated performed marginally better than the LR models built without blood examination variables incorporated in the pre-peak stage.

**Table 10.**
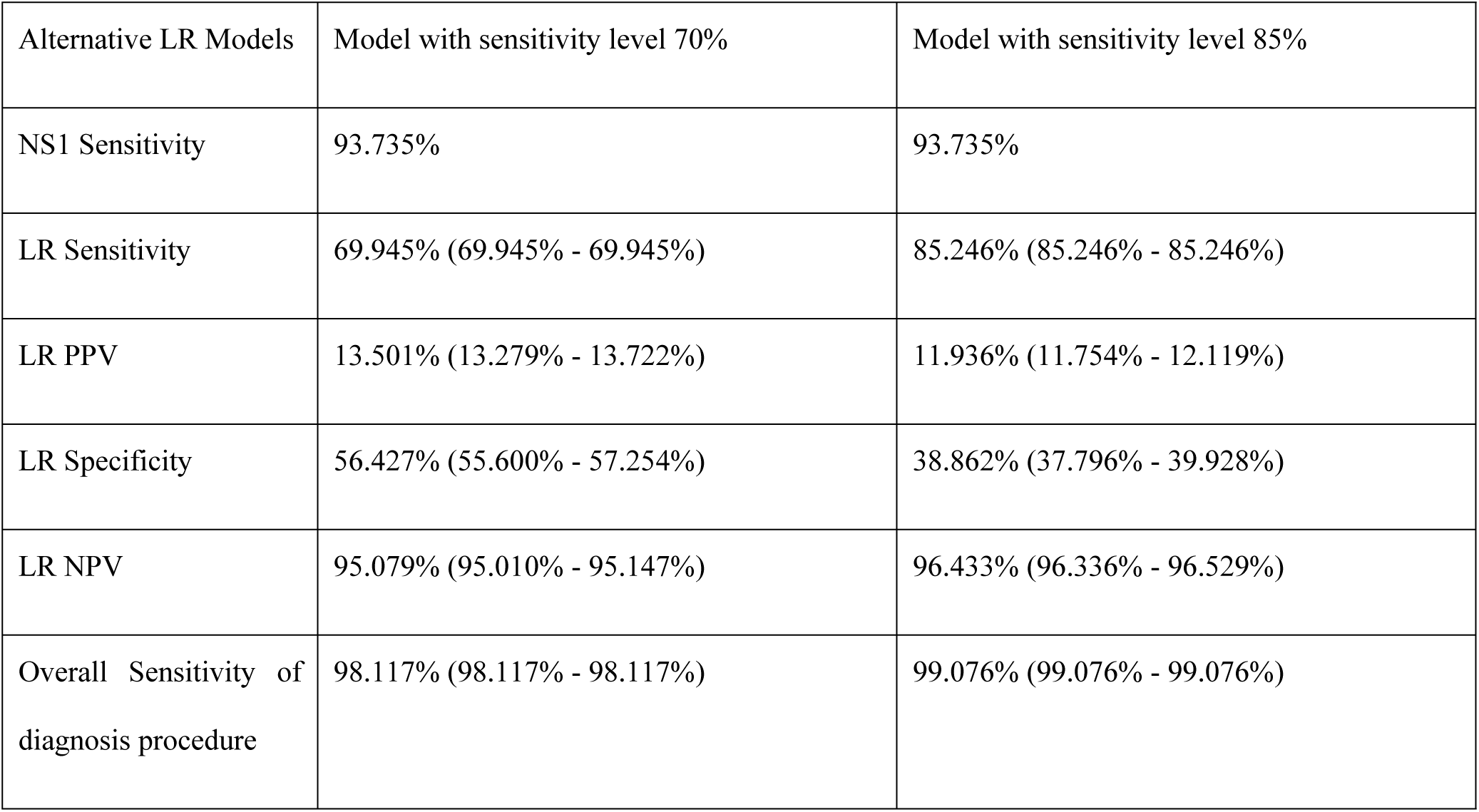
Overall effects of the diagnosis procedure shown in Fig 1 when alternative LR models built without blood examination variables were incorporated. The performance data are reported in terms of mean (95% confidence interval).

**Table 11.** How the LR models built without blood examination variables performed in different stages of the epidemic. The performance data are reported in terms of mean (95% confidence interval).

| Performance metric | Model with sensitivity level 70% | Model with sensitivity level 85% |
| --- | --- | --- |
| RR over the epidemic period | 2.749 (2.666 - 2.832) | 3.365 (3.225 - 3.504) |
| RR_Pre-Peak | 1.626 (1.458 - 1.794) | 2.460 (2.268 - 2.651) |
| RR_Peak | 2.369 (2.268 - 2.470) | 3.240 (3.085 - 3.396) |
| RR_Post-Peak | 5.390 (4.871 - 5.909) | 4.946 (4.322 - 5.570) |
| F1 over the epidemic period | 0.226 (0.223 - 0.229) | 0.209 (0.207 - 0.212) |
| F1_Pre-Peak | 0.281 (0.268 - 0.294) | 0.312 (0.304 - 0.319) |
| F1_Peak | 0.209 (0.205 - 0.212) | 0.201 (0.199 - 0.204) |
| F1_Post-Peak | 0.237 (0.233 - 0.241) | 0.197 (0.194 - 0.201) |
| PPV over the epidemic period | 13.501% (13.279% - 13.722%) | 11.936% (11.754% - 12.119%) |
| PPV_Pre-Peak | 18.908% (17.992% - 19.823%) | 19.471% (18.961% - 19.981%) |
| PPV_Peak | 12.423% (12.182% - 12.664%) | 11.429% (11.257% - 11.601%) |
| PPV_Post-Peak | 13.835% (13.568% - 14.103%) | 11.083% (10.859% - 11.307%) |
| Sensitivity over the epidemic period | 69.945% (69.945% - 69.945%) | 85.246% (85.246% - 85.246%) |
| Sensitivity_Pre-Peak | 54.615% (52.257% - 56.974%) | 78.269% (76.604% - 79.935%) |
| Sensitivity_Peak | 65.319% (64.458% - 66.180%) | 84.043% (83.383% - 84.702%) |
| Sensitivity_Post-Peak | 83.175% (82.106% - 84.243%) | 89.921% (88.983% - 90.859%) |
| Specificity over the epidemic period | 56.427% (55.600% - 57.254%) | 38.862% (37.796% - 39.928%) |
| Specificity_Pre-Peak | 58.750% (57.754% - 59.746%) | 43.041% (41.867% - 44.214%) |
| Specificity_Peak | 57.490% (56.537% - 58.443%) | 39.902% (38.781% - 41.023%) |
| Specificity_Post-Peak | 54.435% (53.709% - 55.161%) | 36.520% (35.434% - 37.606%) |
| NPV over the epidemic period | 95.079% (95.010% - 95.147%) | 96.433% (96.336% - 96.529%) |
| NPV_Pre-Peak | 88.035% (87.366% - 88.703%) | 91.828% (91.136% - 92.521%) |
| NPV_Peak | 94.730% (94.603% - 94.858%) | 96.441% (96.302% - 96.580%) |
| NPV_Post-Peak | 97.353% (97.180% - 97.525%) | 97.615% (97.372% - 97.859%) |

Overall, it is observed that the LR model built with blood examination variables incorporated and with a sensitivity level at 85% delivered a significantly higher RR and a marginally higher F1 score than the alternative prediction models addressed above in the pre-peak stage of the epidemic.

### The CART models

Though the performance data in Tables 6 and 7 reveal that the CART models were marginally inferior to the LR models and the SVM models, the explicit and interpretable prediction rules output by the CART models are of interest to many physicians. Fig 4 and Fig 5 show the prediction rules of the CART models that were built with and without the blood examination variables and delivered a sensitivity level at the 85% level, respectively. One interesting observation is that both models place age at the root and employ the same cutoff. Accordingly, we carried out additional analyses to determine the rationale behind this observation. The data in Table 12 show that the sensitivity level of the NS1 test was higher when applied to patients under 46.5 years old than when applied to patients over 46.5 years old. These numbers suggest that the physicians should not have strong confidence in NS1 results when treating older patients.

**Fig 4.**
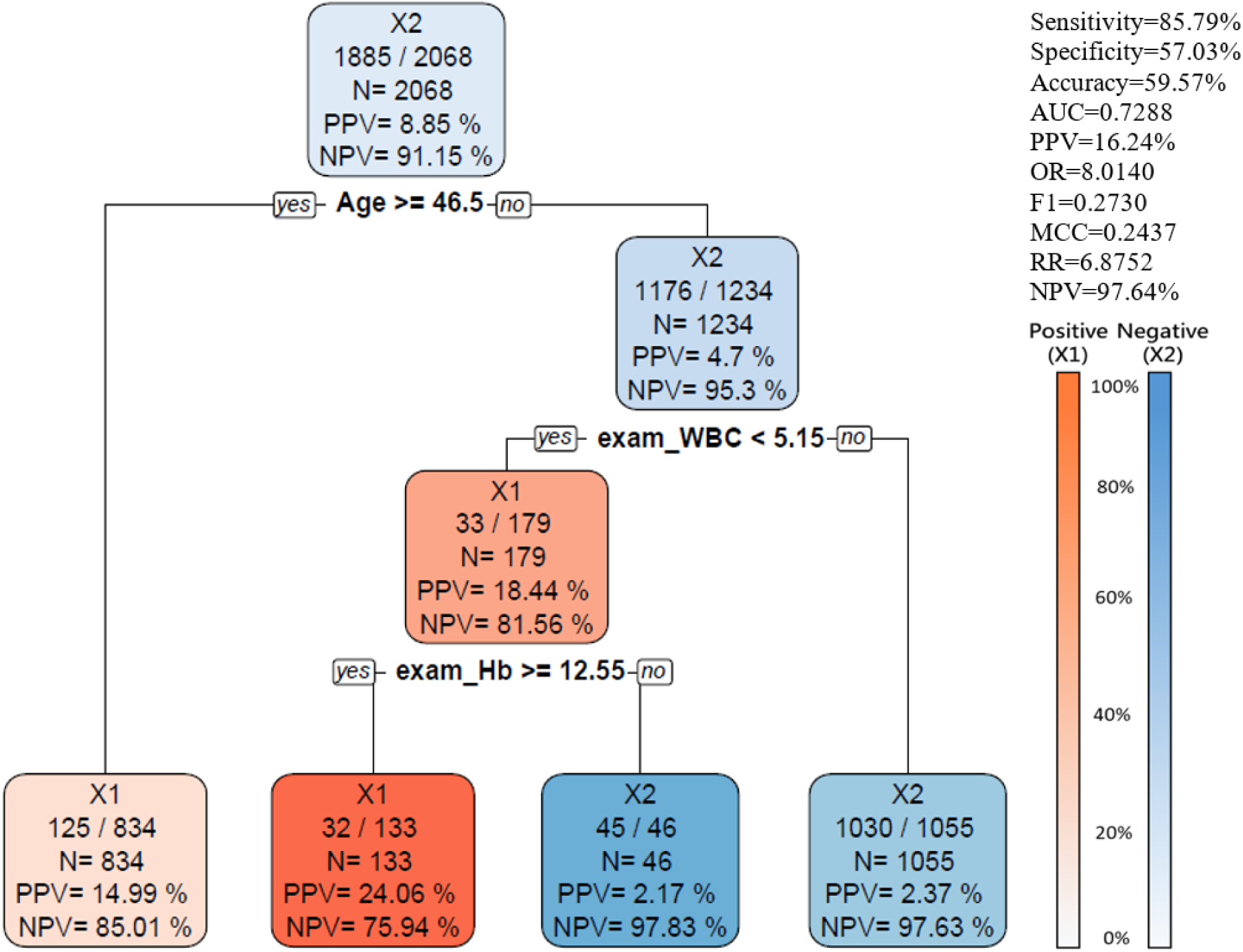
The CART model built with blood examination variables and delivering a sensitivity at the 85% level. Abbreviation: AUC (Area Under the Receiver Operating Characteristics Curve), PPV (Positive Predictive Value), OR (Odds Ratio), NPV (Negative Predictive Value), RR (Relative Risk), MCC (Matthews Correlation Coefficient).

**Fig 5.**
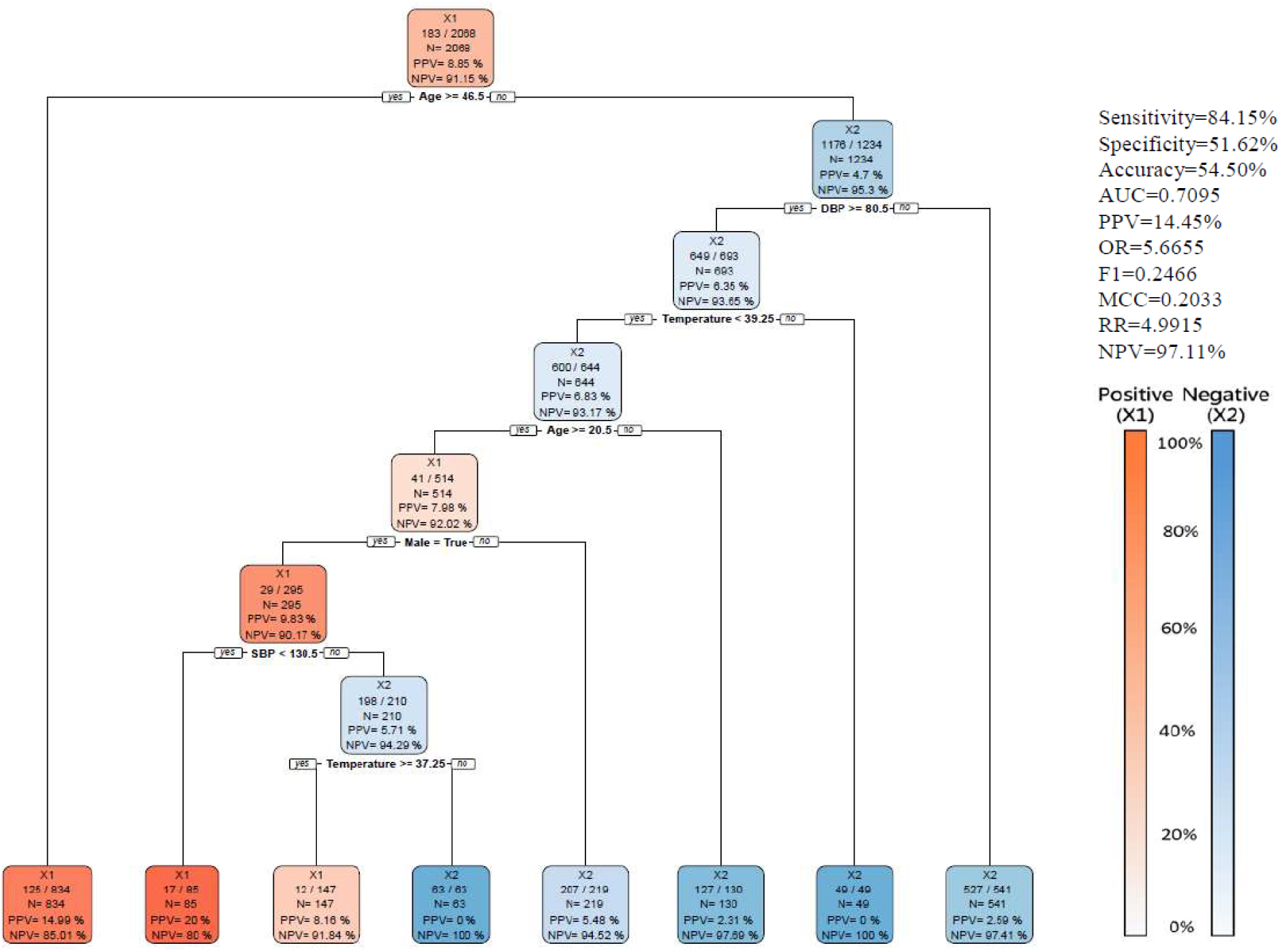
The CART model built without blood examination variables and delivering a sensitivity at the 85% level. Abbreviation: AUC (Area Under the Receiver Operating Characteristics Curve), PPV (Positive Predictive Value), OR (Odds Ratio), NPV (Negative Predictive Value), RR (Relative Risk), MCC (Matthews Correlation Coefficient).

**Table 12.** NS1 sensitivities for DLI patients in different age groups.

|  | Laboratory-confirmed<br>dengue cases with a<br>negative NS1 result | Non-Dengue cases<br>with a negative NS1<br>result | Excluded cases with a<br>positive NS1 result | NS1 Sensitivity |
| --- | --- | --- | --- | --- |
| Emergency visits | 183 | 1885 | 2738 | 93.735% |
| Age (years) |  |  |  |  |
| Age < 46.5 | 58 (31.69%) | 1176 (62.39%) | 1153 (42.11%) | 95.211% |
| Age 46.5 and older | 125 (68.31%) | 709 (37.61%) | 1585 (57.89%) | 92.690% |
| Age (mean±SD) | 55.607±21.144 | 41.798±22.968 | 50.093±21.333 |  |

Abbreviation: SD (standard deviation)

Fig 4 further reveals that the levels of white cell count and hemoglobin were another two important factors to take into account if the patient’s blood examination results were available. On the other hand, Fig 5 reveals that blood pressure levels were other important variables to take into account if the patient’s blood examination results were not available.

## Discussion

In this study, we have demonstrated that machine learning-based prediction models can substantially enhance dengue case detection among patients with a negative NS1 antigen test result. By incorporating routinely collected clinical and laboratory variables into ML frameworks, we achieved a marked improvement in overall diagnostic sensitivity when ML models were integrated with standard NS1 testing. Notably, the experimental results show that by incorporating the logistic regression models calibrated to deliver a sensitivity level of 85%, the overall surveillance sensitivities were raised to over 99%. This high level of sensitivity can effectively mitigate the diagnostic gap associated with NS1 false-negative results [13]. Furthermore, for the suspected dengue patients with a negative NS1 result, the relative risk between those patients that were predicted to be positive by a proposed prediction model and those who were predicted to be negative reached 4.8 when the prediction model was employed in the early stage of a dengue epidemic.

These findings address a critical challenge in dengue control. Although NS1 antigen testing is an indispensable tool for early dengue diagnosis, its imperfect sensitivity—particularly in older adults, secondary infections, and later stages of illness—limits its effectiveness as a standalone screening method. Our results confirm that a non-negligible proportion of laboratory-confirmed dengue cases are NS1-negative and that these cases are clinically meaningful, including individuals with severe outcomes such as ICU admission and mortality. From both clinical and epidemiological perspectives, failure to identify these patients undermines timely care and outbreak containment.

Among the evaluated ML approaches, logistic regression and CART models consistently demonstrated robust and clinically interpretable performance. Logistic regression achieved the highest AUC, while CART models offered transparent decision rules that align closely with clinical reasoning. The interpretability of CART models is particularly valuable for real-world implementation, as it facilitates clinician trust, auditability, and integration into emergency-department workflows.

One important observation on the experimental results is that in the pre-peak stage of the dengue epidemic the LR models built with the blood examination variables incorporated delivered significantly higher sensitivity levels when compared with the LR models built without the blood examination variables and delivering the same levels of overall sensitivity. This finding is clinically intuitive and biologically plausible, as leukopenia, hemoconcentration, and thrombocytopenia are well-recognized hematological features of dengue infection. At the same time, models excluding laboratory variables still performed reasonably well, identifying age, body temperature, blood pressure, and comorbidity profiles as key predictors. This suggests that ML-based screening may remain useful even in resource-limited settings where laboratory testing is delayed or unavailable.

Another important observation is the prominent role of age in CART decision rules. The consistent age cutoff identified by models with and without blood data reflects the reduced sensitivity of NS1 testing in older adults, a phenomenon supported by prior clinical studies. Older patients often present with atypical symptoms and altered immune responses, increasing the likelihood of false-negative antigen tests. Our finding reinforces the need for heightened diagnostic vigilance in this population and suggests that ML-assisted screening be especially beneficial for older adults presenting with dengue-like illness.

From a surveillance standpoint, the performance of ML models varied across epidemic stages, with the greatest net benefit observed during the pre-peak stage. During this stage, ML models achieved higher positive predictive values at comparable sensitivity thresholds, thereby reducing the trade-off cost associated with false-positive alerts. This temporal pattern is particularly relevant for public-health decision-making, as early outbreak detection is disproportionately valuable for initiating vector control and community-level interventions.

Several limitations warrant consideration. First, this study was conducted using data from a single tertiary medical center during a specific outbreak year, which may limit generalizability to other geographic regions, healthcare systems, or dengue serotype distributions. Second, although extensive cross-validation and imbalance-adjustment techniques were employed, external validation using multicenter or prospective datasets is necessary before clinical deployment. Third, the retrospective nature of the analysis precludes assessment of real-time operational impact on clinician behavior and patient outcomes.

Despite these limitations, our study provides compelling evidence that machine learning-assisted screening can meaningfully augment existing dengue diagnostic pathways. By specifically targeting NS1-negative patients—a group traditionally underserved by current diagnostic algorithms—this approach addresses a critical gap at the interface of clinical care and public-health surveillance.

### Clinical and public health implications

Enhanced Sensitivity: Augmenting NS1 antigen testing with ML models substantially increases diagnostic sensitivity for dengue across diverse clinical presentations, improving case detection rates. Triage Support: ML-derived risk scores based on routine clinical data can assist clinicians in stratifying dengue risk and expediting supportive care, especially in high-volume emergency settings. Surveillance Strengthening: Early identification of likely dengue cases—even when NS1 is negative—can improve surveillance accuracy, enabling more timely public health interventions and vector control responses. Resource Optimization: ML models that rely on readily available clinical information are particularly useful where access to PCR is limited, aligning with pragmatic resource constraints in many endemic regions. Targeted Strategies: Age and other demographic factors emerging as important predictors suggest opportunities for stratified screening and follow-up strategies for high-risk groups.

## Author contributions

Conceptualization: Chun-Kai Hwang, Ying-Wen Chen, Tzong-Shiann Ho, Yen-Jen Oyang.

Data curation: Chun-Kai Hwang, Ying-Wen Chen, Tzong-Shiann Ho.

Formal analysis: Chun-Kai Hwang, Ying-Wen Chen, Tzong-Shiann Ho, Yen-Jen Oyang. Funding acquisition: Tzong-Shiann Ho, Yen-Jen Oyang.

Investigation: Chun-Kai Hwang, Ying-Wen Chen, Tzong-Shiann Ho, Yen-Jen Oyang.

Methodology: Chun-Kai Hwang, Ying-Wen Chen, Tzong-Shiann Ho, Yen-Jen Oyang.

Project administration: Tzong-Shiann Ho, Yen-Jen Oyang.

Resources: Chun-Kai Hwang, Ying-Wen Chen, Tzong-Shiann Ho, Yen-Jen Oyang.

Software: Chun-Kai Hwang.

Supervision: Chun-Kai Hwang, Ying-Wen Chen, Tzong-Shiann Ho, Yen-Jen Oyang.

Validation: Chun-Kai Hwang, Ying-Wen Chen, Tzong-Shiann Ho, Yen-Jen Oyang.

Visualization: Chun-Kai Hwang, Ying-Wen Chen, Tzong-Shiann Ho, Yen-Jen Oyang.

Writing – original draft: Chun-Kai Hwang, Ying-Wen Chen, Tzong-Shiann Ho, Yen-Jen Oyang.

Writing – review & editing: Chun-Kai Hwang, Ying-Wen Chen, Yu-Tseng Wang, Tzong-Shiann Ho, Yen-Jen Oyang.

All authors critically revised the manuscript and agreed on the final version.

## Data Availability

The datasets generated and analysed during the current study are not publicly available due to the fact that they constitute an excerpt of research in progress, but are available from the corresponding authors on reasonable request.

## Notes

### Competing Interest Statement

The authors have declared no competing interest.

### Funding Statement

Yes

